# Nutritional Supplementation during Pulmonary Rehabilitation in COPD: A Systematic Review

**DOI:** 10.1101/19000786

**Authors:** Abdulelah M Aldhahir, Ahmed M Al Rajah, Yousef S Aldabayan, Salifu Drammeh, Vanitha Subbu, Jaber S Alqahtani, John R Hurst, Swapna Mandal

## Abstract

**Background:** Uptake of nutritional supplementation during pulmonary rehabilitation (PR) for people with chronic obstructive pulmonary disease (COPD) has been limited by an absence of rigorous evidence-based studies supporting use. Our objective were to report and summarise the current evidence supporting use of nutritional supplementation to improve outcomes during pulmonary rehabilitation in stable COPD patients.

**Methods:** A systematic search was conducted up to May 7^th^, 2019 (registration number CRD42018089142). The Preferred Reporting Items for Systematic Reviews and Meta-Analyses (PRISMA) guidelines were used. Six databases were included: Medical Literature Analysis and Retrieval System Online or MEDLARS Online (Medline), Allied and Complementary Medicine Database (AMED), the Cochrane Database of Systematic Reviews, Excerpta Medica dataBASE (Embase), Cumulative Index of Nursing and Allied Health Literature (CINAHL), and Web of Science.

**Results:** This systematic search generated 580 initial matches, of which 24 studies (1035 COPD participants) met the pre-specified criteria and were included. Our analysis does not confirm an impact of nutritional supplementation during PR, but studies, supplements and PR programmes were heterogeneous in nature.

**Conclusion:** There is currently insufficient evidence on the effect of nutritional supplementation on improving outcomes during PR in patients with COPD. Therefore, controversy remains and further research is needed.

## INTRODUCTION

Patients with COPD tend to have daily symptoms, reduced exercise capacity, and susceptibility to exacerbations, resulting in reduced health-related quality of life.(1-3) The international GOLD strategy document summarises current approaches to COPD management.(1) Cost-effective treatment approaches for COPD, described in the ‘Value Pyramid’ (4) include: smoking cessation, influenza vaccination, and pulmonary rehabilitation. Multiple high-quality randomised controlled trials and meta-analyses have demonstrated that pulmonary rehabilitation is an effective management strategy in COPD patients, since it improves exercise performance, reduces dyspnoea, reduces the risk of exacerbation, and improves health-related quality of life.(5-10)

Exercise intolerance/limitation is one of the most common problems for COPD patients and this may be compounded by reduced muscle mass and malnutrition. It has been reported that COPD patients may lose body weight and skeletal muscle mass, which leads to muscle weakness and dysfunction impacting functional ability and quality of life.(11) Muscle disuse, caused by a prolonged sedentary lifestyle and voluntary immobilisation, leads to muscle deconditioning and thus, reduced muscle strength and endurance.(12) It has also been postulated that COPD is associated with a myopathy, which may be driven by systemic inflammation.(12) Additionally, being underweight is associated with an increased risk of mortality in COPD.(13) Weight loss predicts mortality and morbidity in chronic lung disease patients.(8) Therefore patients with COPD are at risk of significant morbidity and mortality as a result of changes in body composition and nutritional and metabolic status.

It is been suggested that healthy older adults require additional nutrients compared with young adults to preserve bone and lean mass. For instance, it is recommended that young adult require 0.7 g of protein/ kg body weight per day while the recommendation for older adults is 1.2 to 1.5 g protein/kg body weight/day, especially for people with conditions that require higher levels of protein, such as COPD.(14) Nutritional supplements have been used to overcome malnutrition in patients with COPD. It has been suggested that nutritional support integrated with exercise training may improve exercise activity, decrease the risk of mortality, and improved muscle strength in undernourished COPD patients.(15, 16) A meta-analysis of nutritional supplementation for stable chronic obstructive pulmonary disease by Ferreira et al. in 2012 included 17 randomised clinical trials and concluded that nutritional supplements increased muscle mass and body weight, and improved respiratory function and exercise tolerance in COPD patients who were poorly nourished.(17) Additionally, Collins et al. demonstrated in their meta-analysis of nutritional support and functional capacity in chronic obstructive pulmonary disease that nutritional supplements improved weight and handgrip strength in COPD patients.(18) Both reviews only included randomised clinical trials and it was not necessary for participants to be engaged in PR. We hypothesised that an integrated approach of exercise training and nutritional support might be the best way to seek functional improvements. However, uptake of nutritional supplementation **<u>during</u>** pulmonary rehabilitation, where the potential benefit may be greatest, has been limited by the absence of rigorous evidence-based studies supporting use. The objective of this systematic review was to report and summarise the current evidence for using nutritional supplementation during pulmonary rehabilitation in stable COPD patients to enhance PR outcomes.

## METHODS

### Search strategy

The Preferred Reporting Items for Systematic Reviews and Meta-Analyses (PRISMA) guidelines were used for this systematic review, with Prospero registration number CRD42018089142.(19) The search was conducted up to May 7th, 2019 using Medical Literature Analysis and Retrieval System Online or MEDLARS Online (Medline), Excerpta Medica dataBASE (Embase), Allied and Complementary Medicine Database (AMED), the Cochrane Database of Systematic Reviews, Cumulative Index of Nursing and Allied Health Literature (CINAHL), and Web of Science database (table A1, table A2, table A3, table A4, table A5). The search strategy and terms used in this systematic review are described in the Appendix. The bibliography of eligible articles as well as existing systematic reviews in the field were also screened.

### Inclusion criteria

The PICOS (P –population, patient, problem; I –intervention; C –control, comparison or comparator; O –outcome) criteria for included studies appear in Table□1. Studies were included in the systematic review if they met all of the following criteria

**Table 1.**
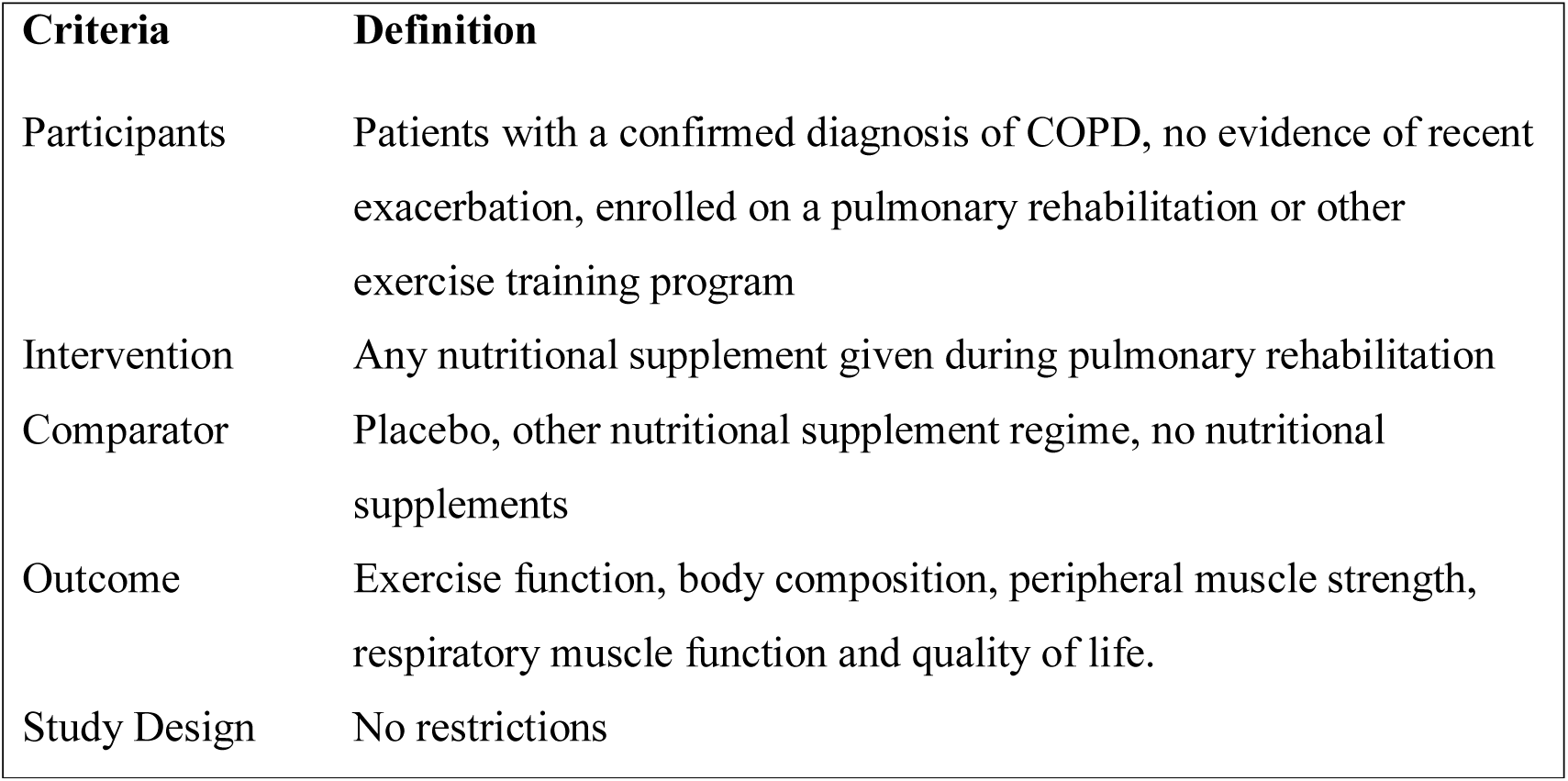
PICOS criteria used for inclusion of studies.

1. Studies of patients with a confirmed diagnosis of COPD.
2. No evidence of recent exacerbation as described in the individual studies.
3. Patients enrolled on a Pulmonary Rehabilitation or other exercise training programme.
4. Patients receiving nutritional supplementation (caloric, non-caloric, powder, liquid, capsule, or tablets) during Pulmonary Rehabilitation or an exercise training program.

### Exclusion criteria

We excluded:

1. Book chapters.
2. Systematic reviews (but screened the reference lists)
3. Non-English manuscripts.
4. Conference abstracts with no full-text.
5. Non-full text articles.

The main outcomes of interest were to investigate the impact of nutritional supplementation during PR programmes on exercise function, body composition, peripheral muscles strength, respiratory muscle function, and quality of life.

### Data collection

Three authors (AA, JH, & SM) screened the titles and abstracts to exclude irrelevant studies. Full texts of the relevant studies were read by the first author (AA) to evaluate if they fulfilled the inclusion criteria. The reference lists of included studies and excluded systematic reviews were also screened; two additional studies were found, and the senior authors (JH & SM) discussed eligibility. Disagreements between authors were resolved by discussion.

### Quality assessment

The first and seventh authors (AA & JH) performed risk of bias assessment using the Cochrane Risk of Bias Tool to assess randomised studies, which comprises seven questions, and the Modified Newcastle-Ottawa scale to assess cohort studies, which is also made up of seven questions.(20, 21) For the randomised trials, we scored each of the seven domains as 0 (low risk of bias) or 1 (high risk of bias, or bias unclear). There was therefore a total score between 0 and 7 in which a higher score equates to a higher risk of bias. For cohort studies, each of the 7 domains was scored from 0 (high risk of bias) to 3 (low risk of bias) and we took a mean of the domains to result in a score between 0 and 3 where a higher score represents a lower risk of bias.

### Synthesis of results

The main purpose of this systematic review was to report and summarise the current evidence of using nutritional supplementation during pulmonary rehabilitation in stable COPD. A meta-analysis was not attempted due to methodological heterogeneity between studies. Our discussion focuses on the studies at lower risk of bias.

## RESULTS

Initially, 580 studies were considered potentially eligible. However, after removing duplicates, 449 titles and abstracts were included. Screening the titles and abstracts resulted in 32 from 449 studies being considered for full-text reading. After reading the full-text of 32 studies, eight further studies were excluded (table A6). Screening the reference list of eligible studies revealed two further relevant studies. Thus 24 studies in total met the inclusion criteria for the systematic review (see Figure 1).

**Figure 1.**
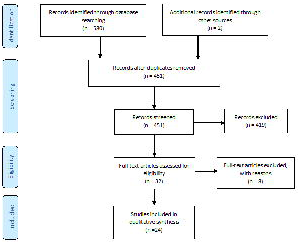
Preferred Reporting Items for Systematic Reviews and Meta-Analyses (PRISMA) Flow Diagram.

The 24 studies comprised five cohort studies and 19 randomised controlled trials (RCTs). The sample size and study duration varied between 8 and 80 participants and six weeks to four months, respectively. A full description of the included RCTs and cohort studies appear in Table 2 and Table 3, respectively. Also, risk of bias assessment for RCT and cohort studies appear in table A7 and table A8, respectively.

**Table 2.** Detailed description of the included RCT studies.

| Author and Risk of Bias | Subject | Intervention | Pulmonary Rehabilitation | Outcomes Measures | Result |
| --- | --- | --- | --- | --- | --- |
| Beers et al.<br>2019<br>(22)<br>**BIAS:**<br>2/7 | N= 73<br>(‘low muscle mass’) | **Intervention:**<br>125mL of 9.4 g protein, 28.1 g carbohydrate and 4.1 g fat, leucine, n-3 PUFA and vitamin D.<br>**Placebo:**<br>Flavoured non-caloric aqueous solution<br>**Phase1:** 3x/day & pulmonary rehab<br>**Phase2** (8 months):<br>1x/day of intervention only. | **Duration:** 4 Months (**phase 1**)<br>**Location:** outpatient.<br>**Session detail:**<br>40 sessions, 2-3 x/week.<br>1- High intensity endurance exercise by cycle ergometry.<br>2- Treadmill walking.<br>3- Progressive resistance exercise of upper and lower body.<br>4- Education session. | 1. Body Composition: Body weight, BMC, ASM, fat mass.<br>2. Muscle Function: Quadriceps strength.<br>3. Exercise Performance: CET by cycle ergometer.<br>4. Quality of Life: HADS, SGRQ, EQ-5D-3L.<br>5. Physical activity: steps | 1. BODY COMPOSITION: Significant differences between groups in body weight change ($1.54 \pm 0.76$ kg, $P = 0.041$)<br>2. MUSCLE FUNCTION: No significant differences between the groups.<br>3. EXERCISE PERFORMANCE: No significant differences between the groups.<br>4. QUALITY OF LIFE: No significant differences between the groups.<br>5. PHYSICAL ACTIVITY: Significant benefit in physical activity ($1030.1 \pm 459.8$ steps/day $P = 0.025$). |
| Ogasawara et al.<br>2018<br>(23)<br>**BIAS:** | N= 45 | **Intervention:**<br>ProSure: high carbohydrate n-3 PUFA enriched.<br>**Placebo:**<br>ENSURE: high carbohydrate without | **Duration:** not specified.<br>**Location:** in patient.<br>**Session detail:**<br>20- 30 min per day consist of:<br>1- Education.<br>2- Breathing methods.<br>3- Conditioning.<br>4- Exercise training. | 1. Body Composition: BMI, LBM, BCM, LBMI, SMI.<br>2. Quality of Life: CAT.<br>3- Breathlessness Scale: MRC.<br>4. Physical activity: steps during hospitalisation. | 1. BODY COMPOSITION: No significant differences between the groups.<br>2. QUALITY OF LIFE: No significant differences between the groups.<br>3. BREATHLESSNESS SCALE: No significant differences between the groups.<br>5. PHYSICAL ACTIVITY: No significant differences between the groups. |
| 4/7 |  | n-3 PUFA.<br>Both products were given once a day for the duration of hospitalisation. |  |  |  |
| Bool et al.<br>(2017)<br>(24)<br>**BIAS:**<br>1/7 | N= 73<br>(‘low muscle mass’) | **Intervention:**<br>125mL of 9.4 g protein, 28.1 g carbohydrate and 4.1 g fat, leucine, n-3 PUFA and vitamin D once/day.<br>**Placebo:**<br>Flavoured non-caloric aqueous solution. | **Duration:** 4 Months<br>**Location:** outpatient.<br>**Session detail:**<br>40 sessions, 2-3 x/week.<br>1- High intensity endurance exercise by cycle ergometry.<br>2- Treadmill walking.<br>3- Progressive resistance exercise of upper and lower body.<br>4- Education session. | 1. Body Composition: Body mass, BMC, SMM, & FM.<br>2. Muscle Function: Quadriceps muscle strength, MIP.<br>3. Exercise Performance: cycle endurance time (CET) & 6MWT.<br>4. Quality of Life: HADS.<br>5. Physical activity: 7 days. | 1. BODY COMPOSITION: significant improvement in body mass ($1.5 \pm 0.6$ kg, $P < 0.05$) & FM ($1.6 \pm 0.5$ kg, $P < 0.01$) in the intervention group.<br>2. MUSCLE FUNCTION: no significant differences between the groups.<br>3. EXERCISE PERFORMANCE: no significant differences between the groups.<br>4. QUALITY OF LIFE. No significant differences between the groups.<br>5. PHYSICAL ACTIVITY: significant benefit in physical activity ($929.5 \pm 459.2$ steps/day $P < 0.05$). |
| Paulin et al.<br>(2016)<br>(25)<br>**BIAS:**<br>1/7 | N= 16 | **Intervention:**<br>B<sub>12</sub> 500 mg/ day for 8 weeks<br>**Placebo:**<br>Maltodextrin. | **Duration:** 8 weeks.<br>**Location:** outpatient.<br>**Session detail:**<br>3 days/week, 40 minutes of: aerobic and resistance exercise. | 1. Cardiopulmonary exercise testing: Incremental or constant-load protocols. | 1. EXERCISE PERFORMANCE: No significant differences between the groups. |
| Ahnfeldt et al. | N= 35 | **Intervention:**<br>Protein Bar (each | **Duration:** 9 weeks.<br>**Location:** outpatient | 1. Muscle Function: lower muscle strength. | 1. MUSCLE FUNCTION: No significant differences between the groups. |
| (2015)<br>(26)<br>**BIAS:**<br>4/7 |  | 134.8kcal of energy, 9.3g protein, 14.6 carbohydrate, 4.2 fat) 2x/day for 9 weeks.<br>**Placebo:**<br>No. | **Session detail:**<br>A- 1 hour 2x/week & home-based 1x/week of:<br>1- Endurance.<br>2- Resistance.<br>3- Interval training.<br>4- Educational class. | 2. Exercise Performance: SWT.<br>3. Quality of Life: SGRQ. | 2. EXERCISE PERFORMANCE:<br>No significant differences between the groups.<br>3. QUALITY OF LIFE:<br>No significant differences between the groups. |
| Gurgun et al.<br>(2013)<br>(27)<br>**BIAS:**<br>2/7 | N= 30 ('wasted') | **Intervention:**<br>250 ml 83.3% carbohydrate, 30% fat, 16.7% proteins, 3x/day.<br>**Placebo:**<br>No. | **Duration:** 8 weeks.<br>**Location:** outpatient.<br>**Session detail:**<br>2x/week 60–80 min/day:<br>A- Education.<br>B- Exercise training include:<br>1- Warm-up & bicycle ergometer for 15 minutes.<br>2- Treadmill (15 minutes).<br>3- Upper & lower extremity strength (5–10 minutes).<br>4- Breathing and relaxation therapies (15–20 minutes each). | 1. Body Composition: Body weight, BMI, FFMI.<br>2. Exercise Performance: 6MWT, ISWT, & ESWT.<br>3. Quality of Life: SGRQ, HADS.<br>4. Breathlessness Scale: MRC & Borg.<br>5. Muscle Size: Quad<sub>CSA</sub>. | 1. BODY COMPOSITION:<br>Significant improvement in weight ($1.1 \pm 0.9$ kg, $P < 0.05$), BMI ($0.2 \pm 1.4$ kg/m<sup>2</sup>, $P < 0.05$), & FFMI ($0.6 \pm 0.5$ kg/m<sup>2</sup>, $P < 0.05$) in intervention group.<br>2. EXERCISE PERFORMANCE:<br>No significant differences between the groups.<br>3. QUALITY OF LIFE:<br>No significant differences between the groups.<br>4. BREATHLESSNESS SCALE:<br>No significant differences between the groups.<br>5. MUSCLE SIZE: significant increase in Quad<sub>CSA</sub> ($2.5 \pm 4.1$ cm<sup>2</sup>, $P < 0.05$) in the intervention group. |
| Hornikx et al.<br>(2012)<br>(28)<br>**BIAS:**<br>3/7 | N= 49 | **Intervention:**<br>vitamin D monthly dosage (100.000 UI cholecalciferol)<br>**Placebo:**<br>Arachidis oleum: 4 ml. | **Duration:** 3 months.<br>**Location:** outpatient.<br>**Session detail:**<br>3x/week 90 minutes training of:<br>1- Cycling.<br>2- Walking on treadmill.<br>3- Stair climbing & Arm cranking.<br>4- Strength exercises for extremities. | 1. Muscle Function: quadriceps strength, MIP & MEP.<br>2. Exercise Performance: incremental cycle ergometer & 6MWD.<br>3. Quality of Life: CRDQ. | 1. MUSCLE FUNCTION:<br>Significant increase in MIP ($11 \pm 12$ cmH<sub>2</sub>O, $P = 0.004$) but no differences between groups in quadriceps strength & MEP.<br>2. EXERCISE PERFORMANCE:<br>No significant differences between the groups.<br>3. QUALITY OF LIFE:<br>No significant differences between the groups. |
| Sugawar a et al.<br>(2012)<br>(29)<br>**BIAS:**<br>1/7 | N= 31 | **Intervention:**<br>MEIN (contains 200kcal 20% protein, 25% lipid, 53.2% sugar, 1.8 fibre, Fisher is 3.7, antioxidant vitamin A&C&E) (2x/day 200 ml) for 12 weeks + provided meal with dietary instruction.<br>**Placebo:**<br>No. | **Duration:** 12 weeks.<br>**Location:** Home-based.<br>**Session detail:**<br>A- Breathing retraining:<br>1- Pursd-lip breathing.<br>2- Diaphragmatic breathing.<br>3- Slow deep breathing.<br>B- Exercise training:<br>1- Upper and lower limb exercises.<br>2- Respiratory muscle stretching calisthenics.<br>3- Level walking for least 15 minutes.<br>4- Inspiratory& expiratory muscle exercises.<br>C- Education program.<br>D- Physiotherapist Supervision every 2 weeks in hospital.<br>E- Periodic visits at home. | 1. Body Composition: Body weight, FFM, FMI, (AC), (AMC), %IBW.<br>2. Muscle Function: MIP & MEP, quadriceps strength<br>3. Exercise Performance: 6MWD.<br>4. Quality of Life: CRQ.<br>5. Breathlessness Scale: MRC. | *Data reported as change in ratio in interventional group vs placebo group, not as absolute values.*<br>1. BODY COMPOSITION: Significant improvement in body weight ($2.6 \pm 3$ kg vs $-0.2 \pm 1.4$ kg, $P = 0.0010$), FMI ($8.6 \pm 10.7$ kg/m<sup>2</sup> vs $0.6 \pm 10.6$ kg/m<sup>2</sup>, $P = 0.048$), %AC ($2.4 \pm 3.7\%$ vs $-0.7 \pm 2.4\%$, $P = 0.0134$), and %IBW ($2.7 \pm 3\%$ vs $-0.2 \pm 1.3\%$, $P = 0.0017$) in the intervention group.<br>2. MUSCLE FUNCTION: MIP ($39.2 \pm 38.9$ cmH<sub>2</sub>O vs $0.1 \pm 24.1$ cmH<sub>2</sub>O, $P = 0.0030$) & quadriceps strength ($10.0 \pm 13.3$ kg/kg vs $-1.6 \pm 9.5$ kg/kg, $P = 0.0079$) increased significantly in the intervention group.<br>3. EXERCISE PERFORMANCE: 6MWD ($19.7 \pm 24.7$ m vs $-7.1 \pm 50.8$ m, $p=0.0137$) improved significantly in the intervention group.<br>4. QUALITY OF LIFE: total score ($6.2 \pm 7.5$ vs $-2.7 \pm 13.1$, $P = 0.0374$) & emotional domain ($8.9 \pm 14.4$ vs $-3.9 \pm 12.2$, $P = 0.0097$) increased significantly in the intervention group.<br>5. BREATHLESSNESS SCALE: MRC $22.6 \pm 40.6$ vs $-4.4 \pm 17.2$ ($P = 0.0339$) improved significantly in the intervention group. |
| Baldi et al.<br>(2010)<br>(30)<br>**BIAS:** | N= 28 depleted. | **Intervention:**<br>Amino acids 4g 2x/day for 12 weeks.<br>**Placebo:**<br>No. | **Duration:** 4 weeks.<br>**Location:** inpatient<br>**Session detail:**<br>5 days/week.<br>30 mins submaximal cycle ergometry.<br>30 minutes walking & 1 arm exercise session. | 1. Body Composition: weight & FFM. | *Data reported as change in interventional group vs change in placebo group.*<br>1. BODY COMPOSITION: Significant increase in weight ($3.8 \pm 2.6$ kg, $P = 0.0002$) vs ($-0.1 \pm 1.1$ kg, $P = 0.81$) and FFM ($1.5 \pm 2.6$ kg, $P = 0.05$) vs ($-0.1 \pm 2.3$ kg, $P = 0.94$). |
| 3/7 |  |  | **THEN:**<br>**Duration:** 8 weeks<br>**Location:** Home<br>**Session detail:**<br>Twice/day 30 minutes unloaded bicycle training. |  |  |
| Laviolette et al.<br>(2010)<br>(31)<br>**BIAS:**<br>2/7 | N= 22. | **Intervention:**<br>Whey protein 20g in 120 ml/day for 16 weeks. (8 without PR and 8 with PR).<br>**Placebo:**<br>Casein 20 g in 120 ml/day for 16 weeks. (8 without PR and 8 with PR). | **Duration:** 8 weeks<br>**Location:** not specified<br>**Session detail:**<br>3x/week. 90mins of:<br>1- Endurance.<br>2- Resistance exercise.<br>3- Education and self-management strategies. | Baseline, 8<sup>th</sup>, & 16<sup>th</sup> week:<br>1. Body Composition: weight<br>2. Muscle Function: quadriceps muscle strength & fatigue<br>3. Exercise Performance: constant work rate cycle endurance<br>4. Quality of Life: CRQ<br>5. Lung Function: spirometry & lung volumes. | 1. BODY COMPOSITION:<br>No significant differences between the groups.<br>2. MUSCLE FUNCTION:<br>No significant differences between the groups.<br>EXERCISE PERFORMANCE:<br>No significant differences between the groups.<br>4. QUALITY OF LIFE:<br>No significant differences between the groups.<br>5. LUNG FUNCTION:<br>No significant difference between groups. |
| Wetering et al.<br>(2010)<br>(32)<br>**BIAS:**<br>3/7 | N= 30 ('wasted') | **Intervention:**<br>Respifor (high-carbohydrate supplement; 125ml, 188 kcal) 3x/day for 4 months.<br>**Placebo:**<br>No. | **Duration:** 4 months.<br>**Location:** outpatient.<br>**Session detail:**<br>1- 2x/week for 30 minutes of intensive exercise.<br>2- 1, 2, 3 months dietician counselling for weight losing and muscle-wasted patients.<br>3- Education program.<br>4- Smoking cessation. | 1. Body Composition: FFMI, BMI.<br>2. Muscle Function: MIP & quadriceps average power.<br>3. Exercise Performance: peak exercise capacity (Wmax), cycle endurance test (CET) and 6MWD.<br>4. Quality of Life: | 1. BODY COMPOSITION:<br>Significant increase in BMI (mean difference 1 kg/m<sup>2</sup>, P < 0.05), and FFMI (mean difference 0.9kg/m<sup>2</sup>, P < 0.05).<br>2. MUSCLE FUNCTION:<br>Significant increase in MIP (mean difference 1.4 kPa, P < 0.05) and QAP (mean difference 13.1 Watt, P < 0.05).<br>3. EXERCISE PERFORMANCE:<br>Significant increase in Wmax (mean difference 11.7 |
| | | | | SGRQ. | Watt, $P < 0.05$), CET (mean difference 525 second, $P < 0.05$), and 6MWD (mean difference 27.2 m, $P < 0.05$).<br>4. QUALITY OF LIFE:<br>No statistically significant difference although absolute difference between groups at 6.1 units is greater than the MCID. |
| Deacon et al.<br>(2008)<br>(33)<br>**BIAS:**<br>2/7 | N=80. | **Intervention:**<br>Creatine<br>**Loading Phase:** 22 g daily, 4 divided doses for 5 days<br>**Maintenance Phase:** (PR) 3.76 g daily.<br>**Placebo:**<br>Lactose. | **Duration:** 7 weeks.<br>**Location:** outpatient<br>**Session detail:**<br>3x/week of:<br>1- Endurance training.<br>2- Individually prescribed resistance training using gym equipment and free weights. | 1. Body Composition: weight, FFM, & FM.<br>2. Muscle Function: quadriceps, triceps, & biceps.<br>3. Exercise Performance: ISWT & ESWT.<br>4. Quality of Life: CRQ-SR. | 1. BODY COMPOSITION:<br>No significant differences between the groups.<br>2. MUSCLE FUNCTION:<br>No significant differences between the groups.<br>3. EXERCISE PERFORMANCE:<br>No significant differences between the groups.<br>4. QUALITY OF LIFE:<br>No significant differences between the groups. |
| Borghi-Silva et al.<br>(2006)<br>(34)<br>**BIAS:**<br>1/7 | N=16. | **Intervention:**<br>Oral L-carnitine 2g, twice/day in 10 ml bottle for 6 weeks.<br>**Placebo:**<br>Saline solution. | **Duration:** 6 weeks.<br>**Location:** outpatient.<br>**Session detail:**<br>1 hour 3x/week:<br>(30 minutes treadmill, inspiratory muscle training). | 1. Body Composition: Triceps skinfold, mid-arm circumference, & BMI.<br>2. Muscle Function: MIP & MEP.<br>3. Exercise Performance: incremental exercise test (treadmill) & 6MWT.<br>4. Breathlessness Scale: Borg. | *Data reported as change in interventional group vs change in placebo group.*<br>1. BODY COMPOSITION:<br>No significant differences between the groups.<br>2. MUSCLE FUNCTION:<br>MIP ($40 \pm 14$ cmH<sub>2</sub>O vs $14 \pm 5$ cmH<sub>2</sub>O, $P < 0.05$) but not MEP, increased significantly in the intervention group.<br>3. EXERCISE PERFORMANCE:<br>No significant differences between the groups.<br>4. BREATHLESSNESS:<br>No significant differences between the groups. |
| Faager et al.<br>(2006)<br>(35)<br>**BIAS:**<br>1/7 | N= 23. | **Intervention:**<br>Creatine 0.3 g/kg body weight/day, divided in 4 doses/day for 7 days.<br>Creatine 0.07 g/kg body weight/day 1 dose/day for remaining 7 weeks.<br>**Placebo:**<br>Glucose. | **Duration:** 8 weeks.<br>**Location:** outpatient.<br>**Session detail:**<br>2x/week for 60-75 minutes of exercise training & education consisting of :<br>1- Ergometer cycling.<br>2- Arm muscle training with dumbbells.<br>3- Rising & getting up from a stool and getting up onto a low stool.<br>4- Thera band exercises for shoulder girdle.<br>5- Thigh muscle training with weight cuffs.<br>6- Abdominal muscle training.<br>7- Flexibility exercises for thorax & adjacent joints. | 1. Body Composition: weight.<br>2. Muscle Function: Grip strength, maximal right knee strength & fatigue.<br>3. Exercise Performance: ESWT.<br>4. Quality of Life: SGRQ.<br>5. Lung Function: spirometry. | 1. BODY COMPOSITION:<br>No significant differences between the groups.<br>2. MUSCLE FUNCTION:<br>No significant differences between the groups.<br>3. EXERCISE PERFORMANCE:<br>No significant differences between the groups.<br>4. QUALITY OF LIFE:<br>No significant differences between the groups.<br>5. LUNG FUNCTION:<br>No significant differences in FEV<sub>1</sub> between the groups. |
| Broekhuizen et al.<br>(2005)<br>(36)<br>**BIAS:**<br>3/7 | N= 80. | **Intervention:**<br>PUFA 1g 9 capsules/day.<br>**Placebo:**<br>9 capsules/day of palm & sunflower oil, vitamin E.<br>**Depleted patients**<br>n=48 Respifor (see above) 3x/day. | **Duration:** 8 weeks.<br>**Location:** inpatient.<br>**Session detail:**<br>A- General physical training of:<br>1- Exercise in relation to daily activities.<br>2- Cycle ergometry.<br>3- Treadmill walking.<br>4- Swimming.<br>B- Sports & games.<br>C- Educational program.<br>D- Regular meals. | 1. Body Composition: BMI, weight, FFM, FM, & FFMI.<br>2. Muscle Function: quadriceps strength, handgrip & MIP<br>3. Exercise Performance: endurance time<br>Incremental bicycle ergometry & Submaximal bicycle ergometry.<br>4. Lung Function: | 1. BODY COMPOSITION:<br>No significant differences between the groups.<br>2. MUSCLE FUNCTION:<br>No significant differences between the groups.<br>3. EXERCISE PERFORMANCE:<br>Maximal exercise capacity (peak workload (9.7 W difference, P = 0.009) & bicycle ergometry duration (4.3 minutes difference, P = 0.023) improved significantly in the intervention group.<br>4. LUNG FUNCTION:<br>No significant differences between the groups. |
|  |  |  |  | spirometry |  |
| Fuld et al.<br>(2005)<br>(37)<br>**BIAS:**<br>3/7 | N= 25. | **Intervention:**<br>Creatine+ Glucose polymer (5g Creatine and 35g glucose/dose).<br>**A-Loading phase:**<br>3x/daily for 14 days.<br>**B-Maintenance phase:**<br>1x/daily for 10 weeks (PR).<br>**Placebo:**<br>Glucose polymer (40.7 g/dose). | **Duration:** 8 weeks<br>**Location:** outpatient<br>**Session detail:**<br>2x/week each 1 hour consisting of:<br>1- A warm-up.<br>2- Mobility training.<br>3- Dynamic strength training of all extremities.<br>4- Whole body endurance training.<br>5- Education and behavioural interventions. | 1. Body Composition: Body mass, FFM, & FM.<br>2. Muscle Function: MIP, lower limb muscle performance, handgrip.<br>3. Exercise Performance: ISWT, ESWT, cycle ergometry.<br>4. Quality of Life: SGRQ.<br>5. Lung Function: Spirometry. | *Data reported as change in interventional group vs change in placebo group.*<br>1. BODY COMPOSITION:<br>FFM increased significantly by (2 kg vs 0.4 kg, $P < 0.05$) in the Creatine group.<br>FM & BM no significant differences between the groups.<br>2. MUSCLE FUNCTION:<br>Significant increase in lower limb strength (19.5 N.m vs 12.2 N.m, $P < 0.05$), endurance (1216 J vs 362 J, $P < 0.05$), handgrip strength (2.9 N vs 0.6 N, $P < 0.05$) & endurance (15.6 repetitions vs 8.4 repetitions, $P < 0.05$) in the Creatine group.<br>No significant change in MIP.<br>3. EXERCISE PERFORMANCE:<br>No significant differences between the groups.<br>4. QUALITY OF LIFE:<br>Total score decreased (5.9, $P < 0.05$) & activity domain decreased (5.3, $P < 0.01$) in the Creatine group.<br>5. LUNG FUNCTION:<br>No significant improvement in FEV<sub>1</sub>. |
| Steiner et al.<br>(2003)<br>(38)<br>**BIAS:**<br>3/7 | N= 60. | **Intervention:**<br>Respifor (high-carbohydrate supplement; 125ml, 188 kcal) 3x/day for 7 weeks<br>**Placebo:**<br>Non-nutritive. | **Duration:** 7 weeks.<br>**Location:** outpatient.<br>**Session detail:**<br>2x/week of:<br>1- Endurance training (walking exercise+ home walking program).<br>2- Circuit of low impact conditioning exercise.<br>3- Educational sessions. | 1. Body Composition: weight, BMI, BM, lean mass, fat mass.<br>2. Muscle Function: quadriceps & handgrip strength.<br>3. Exercise Performance: ISWT& ESWT.<br>4. Quality of Life: | 1. BODY COMPOSITION:<br>Significant improvement in weight (0.63 kg, $P = 0.004$), BMI (0.24 kg/m<sup>2</sup>, $P = 0.002$), & fat mass (0.67 kg, $P = 0.001$) in the intervention group.<br>2. MUSCLE FUNCTION:<br>No significant differences between the groups.<br>3. EXERCISE PERFORMANCE:<br>No significant differences between the groups.<br>4. QUALITY OF LIFE: |
|  |  |  |  | CRQ-SR. | No significant differences between the groups. |
| Vermeer<br>en et al.<br>(2000)<br>(39)<br>**BIAS:**<br>3/7 | Part I:<br>N= 14<br>Part<br>II:<br>N= 11 | **Part I:**<br>**Intervention 1:**<br>1046 kJ, 21%<br>protein, 34% fat,<br>45% carbohydrate.<br>**Intervention 2:**<br>2092 kJ,<br>21%protein, 36%<br>fat, 43%<br>carbohydrate.<br>**Placebo:**<br>209kJ coffee<br>creamers & lemon<br>syrup.<br>**Part II:** (Respifor;<br>see above) vs<br>Pulmocare (high fat<br>supplement) 200 ml. | **Duration:** not specified.<br>**Location:** inpatient.<br>**Session detail:**<br>Not specified. | 1. Exercise Performance:<br>cycle ergometer.<br>2. Lung Function:<br>spirometry.<br>3. Self-Reported:<br>A- Change in<br>breathlessness during<br>meals.<br>B- Leg pain. | 1. EXERCISE PERFORMANCE:<br>**Part I:**<br>No significant differences between the groups.<br>2. LUNG FUNCTION:<br>**Part I:**<br>No significant differences between the groups.<br>**Part II:**<br>PEF (pre 3.1 L/s ± 1.0, post 3.3 L/s ± 1.2) increased<br>significantly after the Respifor supplement vs<br>Pulmocare (pre 3.1 L/s ± 0.9, post 3.1 L/s ± 0.9) (P<br><0.05).<br>3. SELF-REPORTED SYMPTOMS:<br>**Part I:**<br>Satiety changed significantly after the supplements for<br>the 2092-kJ supplement (P < 0.05).<br>**Part II:**<br>Significant increase in breathlessness at 30 and 60<br>minutes following a meal with Pulmocare vs Respifor<br>(raw data not provided, P < 0.05). |
| Schols et<br>al.<br>(1995)<br>(40)<br>**BIAS:**<br>4/7 | N= 71<br>(per<br>protoc<br>ol<br>group)<br>. | Complex, three<br>group study:<br>P group: placebo<br>steroid.<br>N group: placebo<br>steroid + nutritional<br>supplement.<br>N+A: 4 IM<br>injections of<br>nandrolone + | **Duration:** 57 days.<br>**Location:** inpatient.<br>**Session detail:**<br>1- General physical training related<br>to daily activities.<br>2- Cycle ergometry.<br>3- Treadmill walking.<br>4- Walking circuits.<br>5- Swimming. | Measurements were made<br>at entry, 29 and 57 days:<br>1. Body Composition:<br>weight, arm<br>circumference, skinfolds,<br>FFM.<br>2. Muscle Function:<br>MIP.<br>3. Exercise Performance.<br>12MWT. | Comparing group P with group N. Patients were<br>stratified to depleted group vs non-depleted group:<br>**Depleted group:**<br>1. BODY COMPOSITION:<br>No significant difference in FFM or arm circumference<br>between N and P but significant increase in skinfold<br>and weight in the N groups (raw data not provided, P <<br>0.03).<br>**Non-depleted group:**<br>Only reported in per protocol analysis |
|  |  | nutritional supplement (not considered further).<br>Nutrition: 1x/day 200 ml for 57 days mixture of Nutri-drink (high energy), Protifar (high protein) & Fantomalt (high energy carbohydrate), oil). |  |  | 2. MUSCLE FUNCTION:<br>No significant differences between the groups.<br>3. EXERCISE PERFORMANCE:<br>No significant differences between the groups. |

Definition of abbreviation: 12MWT, Twelve-Minute Walk Test; 6MWD, six-minute walk distance; BM, body mass; BMC, bone mineral content; BMI, Body mass index; CRQ, Chronic Respiratory Disease Questionnaire; CWT, constant work rate test; ESWT, Endurance Shuttle Walk Test; FEV<sub>1</sub>, forced expiratory volume in one second; FFM, Fat-free mass; FM, fat mass; FMI, fat mass index; IBW, Ideal body weight; ISWT, Incremental Shuttle Walking Test; MEP, Maximum expiratory pressure; FFMI: fat free mass index; MIP, Maximum inspiratory pressure; MMC, mid-arm muscle circumference; PEF, peak expiratory flow; QAP, quadriceps average power; Quad<sub>CSA</sub>, quadriceps cross-sectional area; SGRQ, St. George's Respiratory Questionnaire; SMM, skeletal muscle mass; UI, International Unit; LBM, Lean body mass LBMI, Lean body mass Index; SMI, skeletal muscle mass index; BCM, Body cell mass; BMC, bone mineral content; ASM, appendicular skeletal muscle mass; EQ-5D-3L, EuroQoL Five-Dimensions Questionnaire.

**Table 3.** Detailed description of the included Cohort studies.

| Author and Risk of Bias | Subject | Intervention | Pulmonary Rehabilitation | Outcomes Measures | Result |
| --- | --- | --- | --- | --- | --- |
| Kubo et al.<br>(2006)<br>(41)<br>**BIAS:**<br>2.4 | N=8. | **Intervention:**<br>400 kcal and 8g protein and abundance of branched chain amino acids in 200 ml.<br>**Placebo:**<br>No. | **Duration:** 8 weeks.<br>**Location:** outpatient.<br>**Session detail:**<br>1x/week for 8 weeks:<br>1- 90 minutes lecture & physical therapy:<br>A- Breathing instruction<br>B- Muscle strengthening exercise for lower limb. | 1. Exercise performance: 6MWD.<br>2. Quality of Life: CRQ. | 1. EXERCISE PERFORMANCE:<br>No significant differences between the groups.<br>2. QUALITY OF LIFE:<br>No significant differences between the groups. |
| Broekhuizen et al.<br>(2005)<br>(42)<br>**BIAS:**<br>1.1 | N= 19<br>Historical Controls: =20. | **Group A:**<br>Respifor (as above) 125ml 3x/day<br>**Group B:** Historical<br>One Ensini (high carbohydrate supplement), one Fortimel (high carbohydrate supplement), one Nutridrink (high carbohydrate supplement), 200 ml 3x/day for 8 weeks. | **Duration:** 8 weeks.<br>**Location:** inpatient.<br>**Session detail:**<br>Daily:<br>1- 2x 20 minutes submaximal cycle ergometry.<br>2- 1x 20 minutes treadmill exercise.<br>3- 1x 30 minutes gymnastics.<br>4- One session of unsupported arm endurance & strength exercise training.<br>5- Educational programme. | 1. Body Composition: weight, FFM, FFMI, and FM.<br>2. Exercise Performance: incremental bicycle ergometry.<br>3. Quality of Life: SGRQ.<br>4. Lung Function: FEV<sub>1</sub>. | 1. BODY COMPOSITION:<br>**Group A:**<br>1- significant weight gain (1.9 kg, P = 0.019) vs group B (1.2 kg)<br>**Both groups:**<br>Post PR, significant gain in weight (A: 1.9 kg, P <0.001; B: 1.2 kg, P < 0.001), FM (only group A 1.3 kg, P < 0.05), and FFM (A: 2kg, P <0.001; B: 1.9 kg, P < 0.05).<br>2. EXERCISE PERFORMANCE:<br>**Both groups:**<br>Peak workload increased significantly during the incremental bicycle ergometry test (Group A: 8.3 ± 17.1 watt, P = 0.062; Group B: 9 ± 9.4 watt, P = 0.002).<br>3. QUALITY OF LIFE:<br>SGRQ<br>**Group A:** |
|  |  |  |  |  | A- No significant differences (although numerical change in SGRQ was greater than the MCID).<br>**Group B:**<br>A- Worse score on the impact dimension.<br>**Both groups:**<br>No significant differences between the groups.<br>4. LUNG FUNCTION:<br>No significant differences between the groups. |
| Creutzberg et al.<br>(2003)<br>(43)<br>**BIAS:**<br>1.7 | N= 64<br>(‘depleted’)<br>Historical Controls = 28. | **Intervention:**<br>Fortimel (as above), Ensini (as above), Fortipudding (high carbohydrate supplement)<br>3x/day for 8 weeks.<br>**Placebo:**<br>No. | **Duration:** 8 weeks.<br>**Location:** inpatient.<br>**Session detail:**<br>A- General physical training:<br>1- Swimming.<br>2- Sports.<br>3- Exercise in relation to daily activities.<br>4- Cycle ergometry.<br>5- Treadmill walking.<br>B- Games.<br>C- Educational program. | 1. Body Composition: weight & FFM.<br>2. Muscle Function: MIP.<br>3. Quality of Life: SGRQ. | 1. BODY COMPOSITION:<br>Significant increase in body weight (2.1 kg, $P < 0.05$) & FFM (1.1 kg, $P < 0.05$) in the intervention group.<br>2. MUSCLE FUNCTION:<br>No significant differences between the groups.<br>3. QUALITY OF LIFE:<br>No significant differences between the groups. |
| Menier et al.<br>(2001)<br>(44)<br>**BIAS:**<br>2.4 | N=60 | **Intervention:**<br>Amino acids 1 capsule/7kg body weight /day, 6 weeks.<br>**Placebo:**<br>No. | **Duration:** 5 weeks.<br>**Location:** not specified<br>**Session detail:** 5 day/week. (40 minutes) Intensity training endurance until exhaustion | 1. Exercise performance: Reached max power (Wmax) | 1. EXERCISE PERFORMANCE:<br>No significant differences between the groups. |
| Creutzberg<br>et al.<br>(2000)<br>(45)<br>**BIAS:**<br>2.2 | N= 24<br>(depleted<br>group). | **Intervention:**<br>Fortimel (as above),<br>Ensini (as above),<br>Fortipudding (as above)<br>3x/day for 8 weeks. | **Duration:** 8 weeks<br>**Location:** inpatient<br>**Session detail:** not specified.<br>Intensity depending on the<br>tolerance of the patient. | 1. Body Composition:<br>weight & FFM. | **Patients divided into (1) no weight gain<2%. (2) expected weight gain >5%. (3) medium weight gain 2 to 5%:**<br>**1. BODY COMPOSITION:**<br>Weight significantly increased for group 3 ($5.8 \pm 1.2$ kg, $P < 0.001$) vs 1 & 2.<br>FFM significantly increased for group 2 (FFM $1.5 \pm 1.2$ kg, $P < 0.05$) & group 3 (FFM $3.1 \pm 1.8$, $P < 0.001$) vs group 1. |

Definition of abbreviation: 12MWT, twelve-minute walk test; 6MWD, six-minute walk distance; BMI, Body mass index; CRQ, Chronic Respiratory Disease Questionnaire; FEV<sub>1</sub>, forced expiratory volume in one second, FFM, Fat-free mass; FFM, Fat-free mass; FFMI: fat free mass index; FM, fat mass; MIP, maximum inspiratory pressure; PR, Pulmonary rehabilitation; SGRQ, St. George's Respiratory Questionnaire.

### Exercise capacity

Data on exercise function, performance, capacity, or endurance were reported in 20 studies using the Endurance Shuttle Walking Test, Incremental Shuttle Walking Test, Six Minute Walk test, Twelve Minute Walk test, treadmill, and incremental or constant work-load cycle ergometry. Seventeen studies found that using nutritional supplements such as high carbohydrates, vitamin D, creatine, or L-carnitine in addition to pulmonary rehabilitation programs had no statistical benefit compared to PR alone.(22, 24-28, 31, 33-35, 37-42, 44) Three studies found that using nutritional supplements (Polyunsaturated fatty acids PUFA and Respifor which is high carbohydrates) had a statistically significant benefit on top of pulmonary rehabilitation.(29, 32, 36)

There was only one study with positive findings at the lowest risk of bias (1/7), in which Sugawara et al. reported increases in Six-Minute Walk Distance (6MWD) by 19.7 ± 24.7 m (less than the minimum clinically important difference). In this RCT the intervention group received a complex supplement twice a day composed of 200 kilocalories, 60% carbohydrates, 15% protein, 25% fat, and 248 μg of omega-3 PUFAs 0.6 with vitamins A, C, and E and a 12 week exercise programme while the control group underwent a 12 week exercise programme only.(29) There were four RCTs with a similarly low risk of bias which demonstrated no benefit of supplementation. Bool et al.(24) reported that using a high carbohydrate supplementation once a day (125 mL of 9.4 g protein, 28.1 g carbohydrate and 4.1 g fat, leucine, n-3 PUFA and vitamin D) over a period of four months within an outpatient pulmonary rehabilitation did not show any significant improvement in exercise performance measured by cycle endurance time (CET) or 6MWT compared to the control PR group who received flavoured non-caloric aqueous solution as a placebo. Similarly, the study by Paulin et al. found that using vitamin B12 for eight weeks during outpatient pulmonary rehabilitation did not show any significant improvement in exercise performance and duration compared to PR alone.(25) Borghi-Silva et al. reported that using L-carnitine twice a day for six weeks did not show significant improvement in exercise performance measured by treadmill and 6MWT when compared to the placebo group, who received saline solution for the same duration.(34) Finally, Faager et al. concluded that using creatine for eight weeks during PR did not improve exercise performance when measured by ESWT compared to the placebo (glucose) group who underwent the same PR.(35)

### Body composition

Seventeen trials measured body composition including body weight, fat-free mass, fat-free mass index, and body mass index.

Body weight was one of the most frequent outcomes measured before and after giving nutritional supplementation; 13 studies measured body weight in COPD patients with normal BMI. Eight studies reported that body weight increased significantly following nutritional supplementation compared to the placebo groups (22, 27, 29, 30, 38, 40, 43, 45), and the study by Broekhuizen et al.(42) compared two nutritional supplements regimes which found that both interventions significantly increased body weight. Four studies reported that body weight did not significantly improve in the intervention groups when compared to the placebo groups.(31, 33, 35, 36) Of the RCTs in which body weight significantly increased, there was only one study, that by Sugawara, that had a low risk of bias.(29) This study reported a significant increase in body weight after 12 weeks of 2.6 ± 3 kg in those receiving the complex supplementation (described above) with mean baseline body weight of 50.8 kg, compared to those in the placebo group with mean baseline body weight of 54.8 kg.(29) In the study by Gurgun et al. there were significant improvements in body weight of 1.1 ± 0.9 kg, BMI 0.2 ± 1.4 kg/m^2^, and in Fat-Free Mass Index (FFMI) (0.6 ±0.5 kg/m^2^) in those who received 250 ml of 83.3% carbohydrate, 30% fat and 16.7% protein three times a day as an intervention.(27) Of the four studies with negative findings, one study was at low risk of bias.(35) This study found no significant difference in body weight between the creatine intervention group and the placebo group after eight weeks.

Body Mass Index (BMI) was assessed before and after using supplementation in six out of 24 studies.(23, 27, 32, 34, 36, 38) BMI significantly increased in the supplementation group when compared to the placebo group in three studies.(27, 32, 38) Three studies reported no significant difference in BMI between participants who received nutritional supplementation with PR compared to PR only.(23, 34, 36) One RCT at the lowest risk of bias showed no improvement in BMI with carnitine.(34) In contrast, Gurgun et al. reported that BMI significantly increased after receiving nutritional supplement.(27)

Fat-free mass (FFM) was evaluated in nine trials.(29, 30, 33, 36, 37, 40, 42, 43, 45) Three studies demonstrated that FFM increased significantly in comparison with the placebo group but these studies all had some risk of bias.(37, 40, 43) Two (27, 32) out of four studies (27, 32, 36, 42) with some risk of bias reported that FFMI significantly increased in the supplemental group when compared to the placebo group. In contrast, the study by Broekhuizen et al. reported no significant difference in FFMI between the group who received PUFA as an intervention and the placebo group who received palm and sunflower oil with vitamin E capsule as a placebo.(36)

### Peripheral muscle strength

Of the 24 studies included in the systematic review, 12 studies measured quadriceps muscles strength, handgrip strength, or both.(22, 24, 26, 28, 29, 31-33, 35-38)

Three studies reported that handgrip strength did not significantly improve in the intervention groups when compared to the placebo groups.(35, 36, 38) Faager et al. being at lowest risk of bias, reported that using carnitine for eight weeks during PR did not significantly improve handgrip strength when compared to the placebo group who received glucose.(35) In contrast, the study by Fuld et al. which had a higher risk of bias, showed significant improvement in the handgrip after using creatine three times a day for two weeks followed by once a day for 10 weeks.(37)

Quadriceps muscle strength was assessed in 12 studies.(22, 24, 26, 28, 29, 31-33, 35-38) Of the 12 RCTs only three studies with 86 participants in total demonstrated positive findings.(29, 32, 37) Sugawara et al. which had a low risk of bias, concluded that quadriceps muscle strength increased significantly after receiving a complex nutritional supplement when compared to the placebo group.(29, 32, 37) However, nine studies reported that using nutritional supplementation during a pulmonary rehabilitation program had no additional effect on quadriceps muscles strength.(22, 24, 26, 28, 31, 33, 35, 36, 38) Bool et al. with a low risk of bias, reported that using a high carbohydrate supplement showed no significant improvement in quadriceps strength when compared to the placebo group.(24) Similarly, the study by Faager et al. showed that using creatine for eight weeks in COPD patients who were enrolled in an eight week PR programme did not reveal significant differences when measuring quadriceps muscles strength compared with those who used placebo.(35)

### Respiratory muscle function

Respiratory muscle function was assessed in nine of the 24 included studies (24, 28, 29, 32, 34, 36, 37, 40, 43), of which three were at lowest risk of bias.(24, 29, 34) Sugawara et al. reported that maximum inspiratory pressure significantly improved in the interventional group (39.2 ± 38.9 cmH_2_O) after receiving the nutritional supplement embedded in 12 weeks of pulmonary rehabilitation compared with the placebo group (0.1 ± 24.1 cmH_2_O).(29) A small study by Borghi-Silva et al. showed a significant improvement in MIP (40 ± 14 cmH_2_O) with carnitine compared to placebo (MIP; 14 ± 5 cmH2O).(34) In contrast, in a larger study by Bool et al. did not show a significant improvement in MIP when compared with the placebo group, who received glucose.(24) None of the studies that measured maximal expiratory pressure showed a significant difference between interventional and placebo groups.(32, 36, 40)

### Quality of life

Quality of life was assessed in 16 out of 24 studies.(22-24, 26-29, 31-33, 35, 37, 38, 41-43) Eight studies used the St. George Respiratory Questionnaire (SGRQ) (22, 26, 27, 32, 35, 37, 42, 43), six used the Chronic Respiratory Questionnaire (CRQ) (28, 29, 31, 33, 38, 41), three used the Hospital Anxiety and Depression Scale (HADS) (22, 24, 27), and only one study used the COPD Assessment Test (CAT). Overall, only two studies demonstrated a significant improvement in quality of life with supplementation in addition to PR.(29, 37) Sugawara et al. which was at lowest risk of bias, quality of life measured by the Chronic Respiratory Disease Questionnaire significantly improved after receiving a nutritional supplement when compared with placebo group.(29) Fourteen studies showed negative findings including two RCTs, at lowest risk of bias, including the study by Faager et al. using creatine supplementation and the study by Bool et al. using the high carbohydrate supplement, which has been describe above. Faager et al. using creatine for eight weeks during PR did not improve quality of life measured by SGRQ.(35) Similarly, Bool et al. reported that four months of using oral nutritional intervention did not improve quality of life measured by HADS.(24)

## DISCUSSION

This review is the first to summarise the potential effects of using nutritional supplementation during pulmonary rehabilitation in patients with COPD. The studies varied in design, and used differing supplements (protein based, vitamin based, amino acid based, carbohydrate based, or fat based), measured various outcomes, and featured different types of pulmonary rehabilitation (home, community, or hospitalised). It is therefore challenging to draw a single conclusion to address whether using a nutritional supplement has additional effects on exercise function, body composition, respiratory muscle function and quality of life during pulmonary rehabilitation. Consequently, appropriately powered studies with suitable designs and sample size to investigate the effect of nutritional support during PR in COPD patients are still needed.

Exercise capacity has been used to quantify the direct effect of nutrition interventions, and to predict mortality and morbidity in COPD patients and other diseases. In this systematic review, the majority of studies demonstrated no improvement in exercise outcomes with nutritional supplementation, compared to PR alone. There were four RCTs with negative findings at low risk of bias (24, 25, 34, 35) which tested carbohydrate, B12, creatine, and carnitine supplementation and just one small RCT with a positive finding which used a complex supplement twice a day composed of 200 kilocalories, 60% carbohydrates, 15% protein, 25% fat, and 248 μg of omega-3 PUFAs 0.6 with vitamins A, C, E.

Body composition is one of the outcome measures that might be expected to improve when using nutritional supplement in COPD patients. Being underweight is associated with an increased risk of mortality in COPD.(13) Low body weight is observed in between 25% and 40% of COPD patients. Among those, 25% have moderate to severe weight loss and 35% have extremely low fat-free mass.(46) In this systematic review, we found that complex nutritional supplementation during PR may increase body weight in population with normal body weight, but we did not find evidence that this occurred with carnitine or creatine. Importantly, improvements in body weight and FFM using nutritional supplementation during pulmonary rehabilitation appear to occur especially in depleted, malnourished, and muscle-wasted patients.(24, 27, 30, 32)

In recent years, researchers have paid attention to the assessment of functional outcomes such as quadriceps muscle strength and handgrip strength. Handgrip strength and quadriceps muscle strength are valid measurements of peripheral muscles strength, and are associated with mortality, morbidity and increased length of hospital stay.(47, 48) In this systematic review, RCTs at low risk of bias did not support the concept that creatine, high carbohydrates, and L-carnitine increase peripheral muscle strength, and we found conflicting evidence for the benefits of complex supplements with one study having positive and another study negative results.

Respiratory muscle weakness in COPD patients may be due to several factors such as acute exacerbations, systemic inflammation, and malnutrition.(49) It has been suggested that nutritional supplements may improve respiratory muscle function. In this systematic review, we found two studies reporting that nutritional supplementation in addition to pulmonary rehabilitation had an extra benefit in improving respiratory muscle function. This was demonstrated by measuring maximum inspiratory and expiratory pressures. The effects were seen only on inspiratory measures, and the authors did not speculate on why they thought this was.

Quality of life may be affected through multiple mechanisms in COPD. The available evidence from this review included one small study demonstrating an improvement in QOL using a complex supplement, and two studies with negative results one of which used creatine and one of which also used a complex supplement.

### Strengths and limitations

To our knowledge, this is the only review that reports the effect of nutritional supplementation **during** pulmonary rehabilitation in stable COPD patients on clinically important outcomes. PR is an evidence-based and cost-effective intervention in COPD and thus maximising outcomes is of great interest to clinicians and patients alike. We have carefully searched the literature and registered our review in advance on PROSPERO. Three independent researchers examined the titles and abstracts for inclusion. Potential limitations are that we only accessed studies in English, and the inherent variation in the included studies, many of which had risk of bias for example with inadequate sample size or absence of a power calculation, variation in outcomes measured, variety in study design, or different pulmonary rehabilitation protocols. It was noticed that there was a variation in the type of supplement either caloric or non-caloric and powder, liquid or tablets. We also observed a variation in the amount, contents and the duration of using supplements.

## CONCLUSION

This is the first systematic review to report the value of nutritional supplementation during PR in patients with COPD. It is not possible to draw a definitive conclusion due to the heterogeneity of the supplements, rehabilitation programmes and outcome measures studied. However, nutritional supplements may enhance the benefit of PR programmes, which would be of considerable benefit to those living with COPD. Not all studies showed positive results and there is a real need for further well-designed and rigorous research to address this area. This is particularly true in weight-losing and/or malnourished patients with COPD who are at the highest risk of poor outcomes.

## Data Availability

Not applicable

## Acknowledgment

We thank Steven Bembridge Medical Librarian at Royal Free London NHS Foundation Trust, UK for his assistance and support in refining the search strategy.

## Contributors

AA, JRH, and SM conceived and designed the study. AA performed the initial search and data extraction, while JRH and SM checked the eligibility of the included articles. AA and JRH performed the quality assessment for the included articles. AA wrote the initial manuscript and YD, JQ, SD, AR contributed to the writing of the manuscript. JRH, SM, VS revised the manuscript. All authors read and approved the final manuscript.

## Funding

Not required.

## Competing interests

JRH, SM, and AA are running a RCT of protein supplementation to enhance PR outcomes in COPD. The product is being supplied by Nutricia. JRH received grants outside the submitted work from pharmaceutical companies that make medicines to treat COPD.

## Patient consent

not required

## Provenance and peer review

Not commissioned; externally peer reviewed.

## Data sharing statement

All data relevant to the study are included in the article or uploaded as supplementary information.

## Appendix

**Table A1.** Medline Search Strategy.

|  |  |  |
| --- | --- | --- |
| 1 | exp Pulmonary Disease, Chronic Obstructive/ | (51369) |
| 2 | (obstruct\$ adj3 (pulmonary or lung\$ or airway\$ or airflow\$ or bronch\$ or respirat\$)).mp. [mp=title, abstract, original title, name of substance word, subject heading word, keyword heading word, protocol supplementary concept word, rare disease supplementary concept word, unique identifier, synonyms] | (110202) |
| 3 | emphysema.mp. [mp=title, abstract, original title, name of substance word, subject heading word, keyword heading word, protocol supplementary concept word, rare disease supplementary concept word, unique identifier, synonyms] | (33167) |
| 4 | (copd or coad or cobd).mp. [mp=title, abstract, original title, name of substance word, subject heading word, keyword heading word, protocol supplementary concept word, rare disease supplementary concept word, unique identifier, synonyms] | (42175) |
| 5 | (chronic adj3 bronchitis).mp. [mp=title, abstract, original title, name of substance word, subject heading word, keyword heading word, protocol supplementary concept word, rare disease supplementary concept word, unique identifier, synonyms] | (11093) |
| 6 | 1 or 2 or 3 or 4 or 5 | (147446) |
| 7 | exp Dietary Supplements/ | (68218) |
| 8 | exp Nutritional Support/ | (43349) |
| 9 | ((diet\$ or food or nutrition\$ or herbal) adj3 (supplement\$ or support\$ or enhance\$)).mp. [mp=title, abstract, original title, name of substance word, subject heading word, keyword heading word, protocol supplementary concept word, rare disease supplementary concept word, unique identifier, synonyms] | (110391) |
| 10 | 7 or 8 or 9 | (158546) |
| 11 | exp Rehabilitation/ | (285709) |
| 12 | rehab\$.mp. [mp=title, abstract, original title, name of substance word, subject heading word, keyword heading word, protocol supplementary concept word, rare disease supplementary concept word, unique identifier, synonyms] | (304313) |
| 13 | 11 or 12 | (497231) |
| 14 | 6 and 10 and 13 | (140) |

**Table A2.** Embase Search Strategy.

|  |  |
| --- | --- |
| 1 exp chronic obstructive lung disease/ | (51369) |
| 2 exp emphysema/ | (14325) |
| 3 exp chronic bronchitis/ | (1712) |
| 4 (obstruct\$ adj3 (pulmonary or lung\$ or airway\$ or airflow\$ or bronch\$ or respirat\$)).mp. [mp=title, abstract, heading word, drug trade name, original title, device manufacturer, drug manufacturer, device trade name, keyword, floating subheading word, candidate term word] | (110202) |
| 5 emphysema.mp. [mp=title, abstract, heading word, drug trade name, original title, device manufacturer, drug manufacturer, device trade name, keyword, floating subheading word, candidate term word] | (33167) |
| 6 (copd or coad or cobd).mp. [mp=title, abstract, heading word, drug trade name, original title, device manufacturer, drug manufacturer, device trade name, keyword, floating subheading word, candidate term word] | (42175) |
| 7 (chronic adj3 bronchitis).mp. [mp=title, abstract, heading word, drug trade name, original title, device manufacturer, drug manufacturer, device trade name, keyword, floating subheading word, candidate term word] | (11093) |
| 8- 1 or 2 or 3 or 4 or 5 or 6 or 7 | (149229) |
| 9 exp diet supplementation/ | (0) |
| 10 exp nutritional support/ | (43349) |
| 11 ((diet\$ or food or nutrition\$ or herbal) adj3 (supplement\$ or support\$ or enhance\$)).mp. [mp=title, abstract, heading word, drug trade name, original title, device manufacturer, drug manufacturer, device trade name, keyword, floating subheading word, candidate term word] | (110391) |
| 12 9 or 10 or 11 | (142817) |
| 13 exp rehabilitation/ | (285709) |
| 14 rehab\$.mp. [mp=title, abstract, heading word, drug trade name, original title, device manufacturer, drug manufacturer, device trade name, keyword, floating subheading word, candidate term word] | (304313) |
| 15- 13 or 14 | (497231) |
| 16- 8 and 12 and 15 | (140) |

**Table A3.** Allied and Complementary Medicine Database Search Strategy.

|  |  |  |
| --- | --- | --- |
| 1 | exp Pulmonary Disease, Chronic Obstructive/ | (48121) |
| 2 | (obstruct\$ adj3 (pulmonary or lung\$ or airway\$ or airflow\$ or bronch\$ or respirat\$)).mp. [mp=title, abstract, original title, name of substance word, subject heading word, keyword heading word, protocol supplementary concept word, rare disease supplementary concept word, unique identifier, synonyms] | (104520) |
| 3 | emphysema.mp. [mp=title, abstract, original title, name of substance word, subject heading word, keyword heading word, protocol supplementary concept word, rare disease supplementary concept word, unique identifier, synonyms] | (32308) |
| 4 | (copd or coad or cobd).mp. [mp=title, abstract, original title, name of substance word, subject heading word, keyword heading word, protocol supplementary concept word, rare disease supplementary concept word, unique identifier, synonyms] | (38816) |
| 5 | (chronic adj3 bronchitis).mp. [mp=title, abstract, original title, name of substance word, subject heading word, keyword heading word, protocol supplementary concept word, rare disease supplementary concept word, unique identifier, synonyms] | (10902) |
| 6 | 1 or 2 or 3 or 4 or 5 | (141003) |
| 7 | exp Dietary Supplements/ | (61794) |
| 8 | exp Nutritional Support/ | (42221) |
| 9 | ((diet\$ or food or nutrition\$ or herbal) adj3 (supplement\$ or support\$ or enhance\$)).mp. [mp=title, abstract, original title, name of substance word, subject heading word, keyword heading word, protocol supplementary concept word, rare disease supplementary concept word, unique identifier, synonyms] | (102371) |
| 10 | 7 or 8 or 9 | (148194) |
| 11 | exp Rehabilitation/ | (272399) |
| 12 | rehab\$.mp. [mp=title, abstract, original title, name of substance word, subject heading word, keyword heading word, protocol supplementary concept word, rare disease supplementary concept word, unique identifier, synonyms] | (170043) |
| 13 | 11 or 12 | (380519) |
| 14 | 6 and 10 and 13 | (125) |
| 15 | pulmonary disease chronic obstructive/ or bronchitis/ or pulmonary emphysema/ or lung diseases obstructive/ | (80818) |
| 16 | (obstruct\$ adj3 (pulmonary or lung\$ or airway\$ or airflow\$ or bronch\$ or respirat\$)).mp. [mp=title, abstract, original title, name of substance word, subject heading word, keyword heading word, protocol supplementary concept word, rare disease supplementary concept word, unique identifier, synonyms] | (104520) |
| 17 | emphysema.mp. [mp=title, abstract, original title, name of substance word, subject heading word, keyword heading word, protocol supplementary concept word, rare disease supplementary concept word, unique identifier, synonyms] | (32308) |
| 18 | (copd or coad or cobd).mp. [mp=title, abstract, original title, name of substance word, subject heading word, keyword heading word, protocol supplementary concept word, rare disease supplementary concept word, unique identifier, synonyms] | (38816) |
| 19 | (chronic adj3 bronchitis).mp. [mp=title, abstract, original title, name of substance word, subject heading word, keyword heading word, protocol supplementary concept word, rare disease supplementary concept word, unique identifier, synonyms] | (10902) |
| 20 | 15 or 16 or 17 or 18 or 19 | (151650) |
| 21 | dietary supplements/ | (46793) |
| 22 | ((diet\$ or food or nutrition\$ or herbal) adj3 (supplement\$ or support\$ or enhance\$)).mp. mp=title, abstract, original title, name of substance word, subject heading word, keyword heading word, protocol supplementary concept word, rare disease supplementary concept word, unique identifier, synonyms] | (102371) |
| 23 | 21 or 22 | (102371) |
| 24 | exp rehabilitation/ | (272399) |
| 25 | rehab\$.mp. [mp=title, abstract, original title, name of substance word, subject heading word, keyword heading word, protocol supplementary concept word, rare disease supplementary concept word, unique identifier, synonyms] | (170043) |
| 26 | 24 or 25 | (380519) |
| 27 | 20 and 23 and 26 | (120) |

**Table A4.** CINHAL.

|  |  |  |
| --- | --- | --- |
| S1 | (MH "Pulmonary Disease, Chronic Obstructive+") |  |
| S2 | (MH "Emphysema+") |  |
| S3 | obstruc* N3 (pulmonary OR lung* OR airway* OR airflow* OR bronch* OR respirat*) |  |
| S4 | emphysema |  |
| S5 | COPD OR COAD OR COBD |  |
| S6 | chronic N3 bronchitis |  |
| S7 | S1 OR S2 OR S3 OR S4 OR S5 OR S6 |  |
| S8 | (MH "Nutritional Support+") |  |
| S9 | (diet* OR food OR nutrition* OR herbal) N3 (supplement* OR support* OR enhance*) |  |
| S10 | S8 OR S9 |  |
| S11 | (MH "Rehabilitation+") |  |
| S12 | rehab* |  |
| S13 | S11 OR S12 |  |
| S14 | S7 AND S10 AND S13 | (52) |

**Table A5.** Web of Science.

|  |  |  |
| --- | --- | --- |
| # 1 | TOPIC: (obstruct* NEAR/3 (pulmonary or lung* or airway* or airflow* or bronch* or respirat*)) | (93750) |
| # 2 | TOPIC: (obstruct* NEAR/3 (pulmonary or lung* or airway* or airflow* or bronch* or respirat*))<br>OR TOPIC: (emphysema) OR TOPIC: (chronic NEAR/3 bronchitis) OR TOPIC: (COPD OR COAD OR COBD) | (135223) |
| # 3 | TOPIC: (obstruct* NEAR/3 (pulmonary or lung* or airway* or airflow* or bronch* or respirat*)) OR TOPIC:<br>(emphysema) OR TOPIC: (chronic NEAR/3 bronchitis) OR TOPIC: (COPD OR COAD OR COBD) AND TOPIC:<br>((diet* or food or nutrition* or herbal) NEAR/3 (supplement* or support* or enhance*)) | (114803) |
| # 4 | TOPIC: (obstruct* NEAR/3 (pulmonary or lung* or airway* or airflow* or bronch* or respirat*))<br>OR TOPIC: (emphysema OR (chronic NEAR/3 bronchitis) OR (COPD OR COAD OR COBD)) | (135223) |
| # 5 | TOPIC: ((obstruct* NEAR/3 (pulmonary or lung* or airway* or airflow* or bronch* or respirat*))<br>OR emphysema OR (chronic NEAR/3 bronchitis) OR (COPD OR COAD OR COBD)) | (135223) |
| # 6 | TOPIC: ((obstruct* NEAR/3 (pulmonary or lung* or airway* or airflow* or bronch* or respirat*))<br>OR emphysema OR (chronic NEAR/3 bronchitis) OR (COPD OR COAD OR COBD)) AND TOPIC:<br>((diet* or food or nutrition* or herbal) NEAR/3 (supplement* or support* or enhance*)) | (491) |
| # 7 | TOPIC: ((obstruct* NEAR/3 (pulmonary or lung* or airway* or airflow* or bronch* or respirat*))<br>OR emphysema OR (chronic NEAR/3 bronchitis) OR (COPD OR COAD OR COBD)) AND TOPIC:<br>((diet* or food or nutrition* or herbal) NEAR/3 (supplement* or support* or enhance*)) AND TOPIC:(rehab*) | (102) |
| # 8 | TOPIC: ((obstruct* NEAR/3 (pulmonary or lung* or airway* or airflow* or bronch* or respirat*))<br>OR emphysema OR (chronic NEAR/3 bronchitis) OR (COPD OR COAD OR COBD)) AND TOPIC:<br>((diet* or food or nutrition* or herbal) NEAR/3 (supplement* or support* or enhance*)) AND TOPIC:(rehab*) | (102) |
| # 9 | TS=(obstruct* NEAR/3 (pulmonary or lung* or airway* or airflow* or bronch* or respirat*)) | (93750) |
| # 10 | TS=(emphysema) | (22729) |
| # 11 | TS=(COPD OR COAD OR COBD) | (57609) |
| # 12 | TS=(chronic NEAR/3 bronchitis) | (9926) |
| # 13 | #12 OR #11 OR #10 OR #9 | (135223) |
| # 14 | TS=((diet* or food or nutrition* or herbal) NEAR/3 | (103028) |
| # 15 | TS=(rehab*) | (205904) |
| # 16 | #15 AND #14 AND #13 | (102) |

**Table A6.**
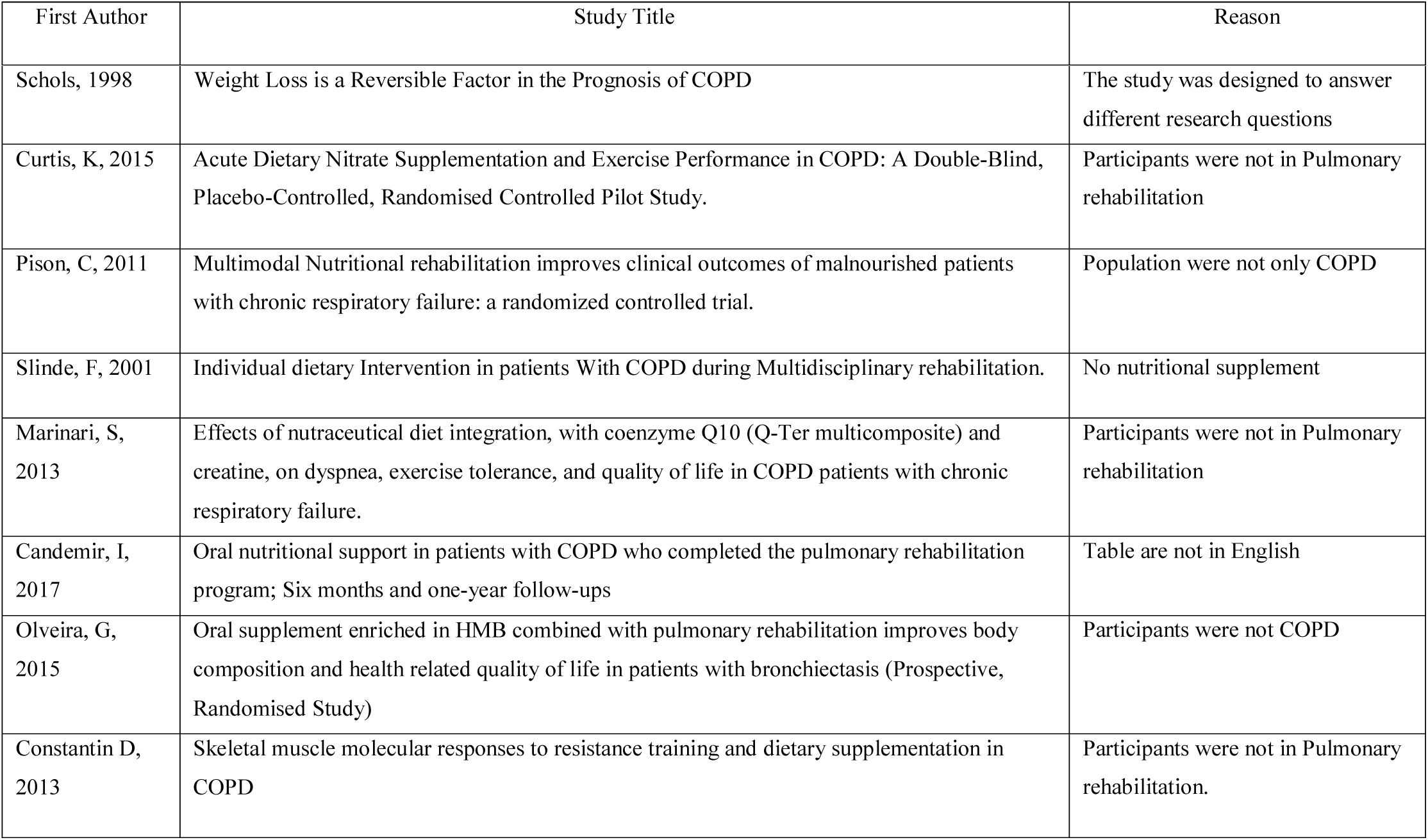
Excluded Studies.

**Table A7.** Risk of bias of the included cohort study.

| First Author | Random Sequence generation | Allocation concealment | Selective reporting | Blinding subject+ personnel | Blinding outcome assessment | incomplete outcome data) | Other source of bias | OVERALL (0-7, higher score = higher risk of bias) |
| --- | --- | --- | --- | --- | --- | --- | --- | --- |
| Bool, 2017 | Low | Low | Low | Low | Low | Low | Unclear | 1 |
| Sugawara, 2012 | Low | Low | Low | Low | Low | Low | Unclear | 1 |
| Paulin, 2016 | Low | Unclear | Low | Low | Low | Low | Low | 1 |
| Faager, 2006 | Low | Unclear | Low | Low | Low | Low | Low | 1 |
| Laviolette, 2010 | Low | Unclear | Low | Low | Low | Low | Unclear | 2 |
| Borghi-Silva, 2006 | Low | Low | Low | Low | High | Low | Low | 1 |
| Gurgun, 2013 | Low | Low | Low | High | Unclear | Low | Low | 2 |
| Beers, 2019 | Low | Unclear | Low | Low | Low | Low | Unclear | 2 |
| Deacon, 2008 | Low | High | Low | Low | Low | Low | Unclear | 2 |
| Vermeeren, 2000 | High | Unclear | Low | Low | Low | Low | Unclear | 3 |
| Baldi, 2010. | Low | Unclear | Low | High | High | Low | Low | 3 |
| Fuld, 2005 | Low | Unclear | Low | Low | Low | High | High( design) | 3 |
| Wetering, 2009 | Low | Low | Low | High | Low | High | Unclear | 3 |
| Broekhuizen, 2005 | Low | High | High | Low | Low | Low | Unclear | 3 |
| Steiner, 2003 | Low | High | Low | Low | Low | High(drop rate) | Unclear | 3 |
| Schols, 1995 | Low | Unclear | Low | High | High | Low | unclear | 4 |
| Hornikx, 2012 | High | High | Low | Low | Low | Low | High | 3 |
| Ogasawara, 2018 | Low | Low | High | High | High | Low | Unclear | 4 |
| Ahnfeldt, 2015 | Low | Low | Low | High | High | High | Unclear | 4 |

**Table A8.** Risk of bias of the included cohort study.

| First author | Population representative | Sample size adequate | Confounders | Statistical analysis | Missing data | Methodology of the outcome | Objective assessment | OVERALL (0-3, higher score = lower risk of bias) |
| --- | --- | --- | --- | --- | --- | --- | --- | --- |
| Creutzberg, 2000 | 3 | 0 | 2 | 3 | 3 | 3 | 3 | 2.4 |
| Broekhuizen, 2005 | 3 | 2 | 0 | 3 | 3 | 3 | 2 | 2.3 |
| Creutzberg, 2003 | 3 | 2 | 0 | 3 | 3 | 3 | 3 | 2.4 |
| Menier, 2001 | 3 | 2 | 0 | 0 | 3 | 1 | 3 | 1.7 |
| Kubo, 2006 | 3 | 0 | 0 | 0 | 3 | 0 | 2 | 1.1 |

0 = definitely no (high risk of bias); 1 = mostly no; 2 = Mostly yes; 3 = definitely yes (low risk of bias)

## Notes

### Competing Interest Statement

Dr. John Hurst, Dr. Swapna Mandal and Abdulelah Aldhahir report grants from Nutricia to run an RCT of protein supplementation to enhance pulmonary rehabilitation outcomes in COPD.
Subbu declared no competing interest
Al Rajeh declared no competing interest
Aldabayan declared no competing interest
Alqahtani declared no competing interest
Drammeh declared no competing interest

### Clinical Trial

This systematic review was registered at Prospero with registration number CRD42018089142

### Funding Statement

This work does not required funding

### Author Declarations

All relevant ethical guidelines have been followed and any necessary IRB and/or ethics committee approvals have been obtained.

Any clinical trials involved have been registered with an ICMJE-approved registry such as ClinicalTrials.gov and the trial ID is included in the manuscript.

